# Development of a clinical decision-support tool for Management of Adolescent knee Pain (The MAP-Knee Tool)

**DOI:** 10.1101/2023.01.11.23284426

**Authors:** Henrik Riel, Malene Kjær Bruun, Chris Djurtoft, Martin Bach Jensen, Søren Kaalund, Guido van Leeuwen, Charlotte Overgaard, Ole Rahbek, Michael Skovdal Rathleff

**Author notes:** Corresponding author Henrik Riel, PhD, Department of Health Science and Technology, Faculty of Medicine, Aalborg University, Selma Lagerløfs Vej 249, 9260 Gistrup.

## Abstract

**Objective:** This study aimed to develop a clinical decision-support tool (The MAP-Knee Tool) to improve the management of adolescents with non-traumatic knee pain.

**Methods:** This multi-step study consisted of five steps ((1-4) initial development and (5) end-user testing with adolescents with or without non-traumatic knee pain and medical doctors). It ended with the first version of the MAP-Knee Tool for the six most common non-traumatic knee pain conditions. The tool includes four components: 1) tool for diagnosing, 2) credible explanations of the diagnoses based on two systematic literature searches and an Argumentative Delphi process with international experts, 3) prognostic factors based on an individual participant data meta-analysis, and 4) option grid including an unbiased presentation of management options based on the available evidence.

**Results:** We included seven children/adolescents (8-15 years old) and seven medical doctors for the end-user testing. All four components were revised accordingly, and the text was condensed as the initial draft was too comprehensive.

**Conclusion:** We developed a clinical decision-support tool for clinicians and adolescents with non-traumatic knee pain to support the consultation in clinical practice.

**Practice Implications:** The tool targets clinicians and adolescents with four components that may decrease diagnostic uncertainty and increase shared decision-making.

## 1. Introduction

Shared decision-making in clinical practice is a complex interpersonal and collaborative process where patients and healthcare professionals make informed decisions about managing the patient’s health.(1) Ideally, decisions are made collaboratively. Information is presented objectively and non-biased, typically in situations where the personal circumstances of patients and their families play a significant role in decisions.(2,3) However, there is a lack of evidence regarding how to increase shared decision-making among patients and clinicians.(1) One method to facilitate a shared decision-making process is using patient decision aids to support the patients’ dialogue with the clinician.(4,5) Decision aids may come in many forms, such as leaflets, videos, or technology-based applications. The common denominator for these aids is that they present the patient with a non-biased and evidence-based overview of different options they may consider and not recommend one over the other.(5,6)

Patient decision aids can support patients in making informed choices regarding their health condition. However, these aids may also indirectly support clinicians in their work while discussing, e.g. treatment options with patients.(7) (Guldhammer et al. *in press)* Such aids may be especially relevant in conditions with several treatment options available.(4,8) An area with multiple treatment options available is musculoskeletal conditions. These conditions are characterised by numerous treatment options that each come with its own set of benefits and harms and time requirements needed from the patient.

Annually, 7% of adolescents visit their general practitioner due to musculoskeletal pain.(9) Especially knee and back pain are prevalent among adolescents, with the majority of knee pain in adolescents having a non-traumatic origin.(10–13) More than one in four adolescents having knee pain will receive care in the secondary or tertiary sectors.(14) Adolescent knee pain was historically viewed as a self-limiting condition. Still, it may severely impact health-related quality of life and physical activity, and almost half of adolescents may continue to experience pain into adulthood.(14–16) This emphasises a need for better management for adolescents with knee pain to reduce recovery times. Results from our previous randomised trial showed that high-cost treatment options such as supervised, physiotherapy-led exercises are only slightly more beneficial than patient education (number needed to treat of 11).(17) However, because of insufficient evidence, guideline recommendations are unclear about the clinical selection of patients who are likely to benefit from referral to additional interventions. Furthermore, the current care pathways of adolescents with non-traumatic knee pain are heterogeneous.(18,19) Therefore, resources may be wasted among adolescents with a good prognosis, while adolescents with a poorer prognosis may not receive sufficient care.

Stratified care based on prognostic factors has been suggested as a possible solution to individualise treatments, as referral to comprehensive rehabilitation may only be needed for some.(20) Yet, for adolescents having non-traumatic knee pain, stratified care based on prognostic factors is currently unavailable. A decision aid including both tools to stratify care and support shared decision-making may help solve some of the practical challenges by supporting both clinicians and patients. This has the potential to minimise unnecessary high-cost referrals and provide higher-value care.

We recently developed a support tool for diagnosing the most common types of non-traumatic adolescent knee pain.(7) This tool improved the diagnostic accuracy of medical doctors.(7) However, there is currently no tool available to support the entire consultation and shared decision-making process when an adolescent suffering from non-traumatic knee pain presents at clinical practice.

The purpose of this study was to develop a decision-support tool to support medical doctors, physiotherapists, and adolescents with non-traumatic knee pain in the shared decision-making process during the entire consultation from diagnosis to deciding on the future management strategy of the adolescents’ condition.

## 2. Material and methods

### 2.1. Study design

This multi-step study consisted of five overarching steps (initial development of the components of the tool and end-user testing), which ended with the first version of a tool to support clinicians and adolescents with the most common non-traumatic knee pain conditions (Patellofemoral Pain, Osgood-Schlatter disease, Patellar Tendinopathy, Sinding-Larsen-Johansson, Growth Pain, and Iliotibial Band Syndrome).(7,17,21,22) The tool was developed to live up to the criteria of the International Patient Decision Aids Standard (IPDAS).(23,24). The study was conducted at the Center for General Practice at Aalborg University and the Department of Health Science and Technology, Faculty of Medicine, Aalborg University.

### 2.2. Development process

The tool was designed to support the entire consultation, from diagnosing the condition to deciding on future management. We included four separate components in the MAP-Knee Tool elements: 1) a tool for diagnosing the most common types of non-traumatic knee pain (SMILE), 2) credible explanations of the aetiology and pathogenesis specific to the diagnosis based on multiple methods with iterative design, 3) a presentation of prognostic factors based on an individual participant data meta-analysis(25), and 4) an option grid that presents the users of the tool with pros and cons of commonly used management options based on a systematic literature search of systematic and narrative reviews within non-traumatic adolescent knee pain. The tool was developed in printed form with two pages including SMILE and the prognostic factors for the clinician exclusively and two pages including the credible explanations and the option grid that the clinician should use when talking to the adolescent and for adolescents to bring home after the consultation. To increase readability and comprehension of the text directed at adolescents, we used a readability assessment tool (the Lesbarkeitsindex (LIX)) to limit the number of complex and lengthy words.

An overarching focus of integrating the four components was to support shared decision-making and base decisions on all three pillars of evidence-based medicine: patient values, clinical expertise, and relevant research.(26) Therefore, the tool should not provide the users with definitive answers simply based on available evidence. The choice of components was based on the natural flow during the consultation, which starts with diagnosing and ends with a decision on the treatment strategy. Before designing the components, a logic model was made inspired by the Implementation Research Logic Model used in implementation research.(27) The authors discussed how each component should affect both adolescents and clinicians, ultimately leading to better treatment and fewer wasted healthcare resources. The logic model underlying the design of the tool can be seen in Figure 1.

**Figure 1:**
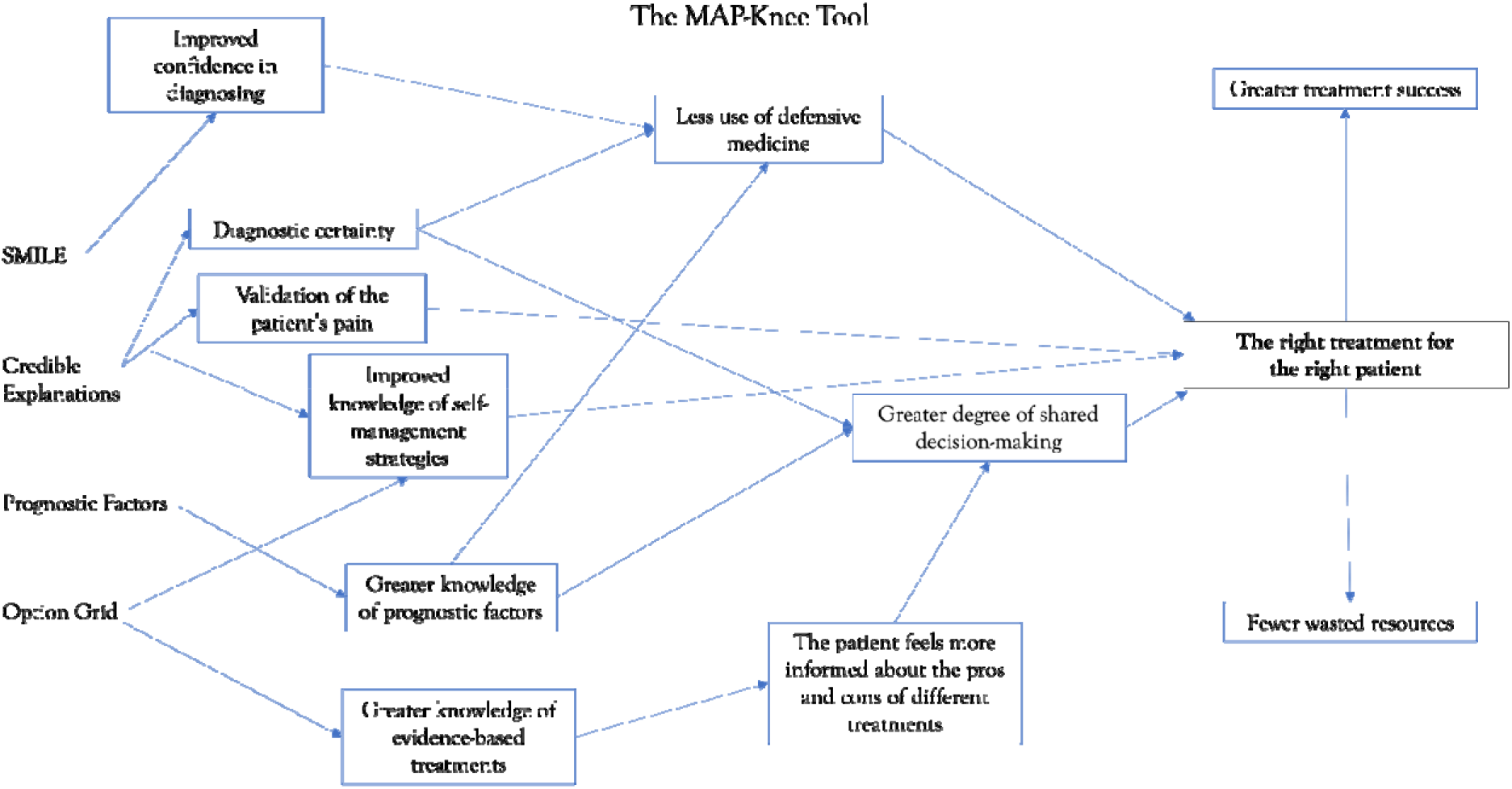
Logic model of the four components of the MAP-Knee Tool with their proposed mechanisms and outcomes.

### 2.3. SMILE

SMILE is previously validated and described in detail elsewhere,(7) whereas the other components are described below.

### 2.4. Credible explanations

The credible explanations were developed using an iterative process of three steps, and the complete methods are published elsewhere.(28) This consisted of 1) two systematic searches of qualitative and quantitative literature (see supplementary file xx) that aimed to answer the *“what information needs to be included in a credible explanation?”*, 2) a two-round Argumentative Delphi process to explore *“what do expert clinicians consider important in a credible explanation”* among clinicians, researchers, and psychologists, and 3) think-aloud exercises with end-users (children, adolescents, and medical doctors). Based on themes from the synthesis of the qualitative literature search and the Argumentative Delphi process, including 16 international experts, we identified three key domains to consider when tailoring credible explanations to adolescents experiencing chronic non-traumatic knee pain. The three key domains were “What is (*diagnosis*) and what does it mean?”, “What is causing my knee pain” and “How do I manage my knee pain?”.(28)

### 2.5. Prognostic factors

The prognostic factors component provided the clinician with knowledge regarding five factors associated with long-term pain and functional deficits in adolescents with non-traumatic knee pain. These factors are lower health-related quality of life, daily or weekly knee pain frequency, knee pain duration over 12 months, bilateral knee pain, and female sex.(25) Three prototypes of this component with different text and graphics were developed by a young medical doctor and PhD student and were individually presented to two medical doctors; one general practitioner with 14 years of experience and one in training to become a general practitioner. The doctors were asked to provide feedback on the three prototypes during single-person interviews and to decide on which prototype they preferred. Inspired by the method used in future workshops, we asked the doctors to critique the prototypes and formulate ideas for improvement.(29)

The three prototypes varied in the degree of controlling the clinician’s advice to the patient and parents based on the number of prognostic factors present. Prototype 1 did not indicate future management. Prototype 2 gave some degree of guidance on future management strategies, including a horizontal bar with numbers from 0 to 5 to indicate the number of prognostic factors present over colour that shifts from green to red to predict poorer prognosis with an increasing number of present prognostic factors. Prototype 3 clearly directed the clinician toward future management of the condition based on a cut-off of prognostic factors present.

### 2.6. Option grid

The purpose of the option grid was to give adolescents, parents, and medical doctors an unbiased presentation of the pros and cons of different treatment strategies. During the development, we used the steps described by Marrin et al. for creating option grids.(30) We applied the same systematic search strategy as previously used for developing the credible explanations, but this time we only included systematic or narrative reviews that compared one or more treatments for either of the six non-traumatic knee pain conditions. Two researchers screened the titles and abstracts and decided which reviews to include. The most recent reviews within each diagnosis and the inclusion of commonly used treatment strategies were prioritised. As a framework for how to visually present the option grid and the recently asked questions patients may pose, we found inspiration in the work by Barr et al.(31) However, to better mimic a clinical guideline considering our setting, we substituted the row with information regarding costs with a clinical recommendation about what will likely be beneficial under which circumstances. We decided by consensus in the author group which treatment categories to include (i.e., which treatments should have a column of their own and which should be included in the ‘Other treatments’ column). These were further sub-grouped based on whether they would require a referral from the orthopaedic surgeon. We prioritised treatments commonly used in the orthopaedic setting as this is the setting in which the tool’s effectiveness will first be investigated and treatments with the best available evidence.

### 2.7. End-user testing

After finalising an initial prototype of the tool and the four components, we performed end-user testing using think-aloud sessions with adolescents having non-traumatic knee pain, adolescents with no history of knee pain, and medical doctors. There was considerable overlap between the tools developed for each diagnosis, so we only used the tool created for Osgood-Schlatter disease during the end-user testing. We applied the feedback to the other diagnoses.

The end-user testing was conducted face-to-face or online via Microsoft Teams, where all information and consent forms were sent by mail for online participation. As the overarching aim of the credible explanation and option grid components was to facilitate meaningful exchanges of information about adolescents’ health status during consultations, we included children and adolescents with and without knee pain and medical doctors with experience treating adolescents with knee pain.

Participants were sampled purposefully based on perceived information power (32), with particular attention to Faulkner’s observations on how 5-8 users will identify 85-95% of all usability problems based on the law of diminishing returns (33). We aimed to include at least one child under the age of the target group (<10 years) to ensure the comprehensibility of the text. All participants were initially instructed in the study procedure and then signed informed consent. The parents or legal guardians signed informed consent on behalf of the child and adolescents under 18 years of age.

All included participants partook in a usability testing of the MAP-Knee Tool based upon the ‘think-aloud approach’ (34). The think-aloud approach was chosen because it focuses on using simulation to gain insights into participants’ interpretations, interactions, reasoning, and latent challenges related to artefact use (35) and using this knowledge to optimise the design. We introduced the complete MAP-Knee Tool to the medical doctors and the credible explanations and option grid to the adolescents. However, doctors’ input regarding credible explanations and the option grid was only used to assess the set-up and layout. Comprehension and content were left to the adolescents. Participants were instructed to verbalise thoughts as they occurred while completing the assigned task and say whatever came to their minds as they went through the MAP-Knee Tool. The researcher would then explore key reflections to identify any misunderstanding or confusion of relevant items and domains. Participants were encouraged to speak constantly as if they were alone in the room and were informed that the researcher would remind them to keep talking should they fall silent.(34) This process was conducted to identify any inconsistencies in the participants’ perception of the items compared to what they were intended to capture (e.g., shared decision-making and diagnostic uncertainty). In addition, this allowed the identification of difficulties in understanding phrases or concepts.

All interviews were recorded and transcribed (non-verbatim). For three reasons, we chose non-verbatim. First, we wanted to make sure that there were no comprehension problems; second, we did not plan to do an in-depth thematic analysis as this would not be needed to identify difficulties in the understanding of the tool; and third, we thought that cutting out all extraneous speech would make the transcript easier to read and more helpful so that we could implement the suggestions made by our participants.(36)

## 3. Results

We included seven Danish children and adolescents aged between 8 and 15 years (three in Session 1 and four in Session 2) and seven medical doctors (three general practitioners, three doctors working in general practice or at the orthopaedic department as part of their clinical education programme, one orthopaedic surgeon) (four in Session 1 and three in Session 2) for the end-user testing. One adolescent in each session suffered from non-traumatic knee pain.

### 3.1. SMILE

As SMILE had previously been validated, no significant changes were made to the content. During the end-user testing, we discovered two misspellings we corrected. To not exclude any sub-groups of patients, we changed the word ‘athletes’ found in the box concerning Sinding-Larsen-Johansson to ‘children and adolescents’. To make the English and Danish versions more consistent, we removed the word ‘pain-free’ from the description of the swelling that could indicate inflammatory arthritis or infection.

### 3.2. Prognostic factors

Prototype 2 was the preferred prototype by both medical doctors during the initial presentation of the three prototypes. They suggested the addition of overlapping brackets below the coloured horizontal bar to indicate either a more conservative management with a lower number of prognostic factors or the consideration of referral to rehabilitation with more prognostic factors. The overlap of the brackets could support clinicians in considering their clinical reasoning rather than simply basing their recommendation of management on a cut-off of the number of prognostic factors present, which was used in Prototype 3.

Most suggested changes during the end-user testing were related to the graphical presentation, not the content. The medical doctors would prefer that the factors were listed on top of each other rather than side by side and that there would be a single arrow below the bar instead of the overlapping brackets. This arrow should have text to explain that more factors would be associated with a poorer prognosis and that more extensive treatment should be considered. They would also prefer to have a text box explaining how to evaluate health-related quality of life in a patient. Therefore, we included a text box with questions the clinician should ask themselves regarding the physical, mental, and social limitations that their patient experiences.

### 3.3. Option grid

We designed the option grid based on information from 20 systematic or narrative reviews (see supplementary file 1) (Osgood-Schlatter disease: n=5, growth pain: n=5, patellar tendinopathy: n=3, patellofemoral pain: n=3, iliotibial band syndrome: n=3, and Sinding-Larsen-Johansson: n=1). Treatments were divided into 1) Watch and wait, 2) Information about knee pain and how to self-manage, 3) Physical activity or exercise, and 4) Passive treatments. After the first draft had been presented to the participants during the end-user testing, we limited the text as it was too elaborate. The medical doctors suggested that ‘Watch and wait’ should be changed to ‘Wait and see’ as this was the term used in everyday practice despite the approach including more than simply waiting. The treatments should be listed in an order that reflects how comprehensive they are. ‘Passive treatments’ should be changed to ‘Other treatments’ as the word ‘passive’ could be interpreted as something negative. Lastly, implementing colour-divided boxes was important to make the table more manageable.

## 4. Discussion and Conclusion

### 4.1. Discussion

We systematically developed a decision support tool for the six most common non-traumatic knee pains in adolescents. The development included both systematic literature searches, an Argumentative Delphi process, and end-user involvement, which ultimately led to the MAP-Knee Tool, which clinicians may use during the consultation with adolescents having non-traumatic knee pain.

Despite the seminal work by The International Patient Decision Aid Standards (IPDAS) Collaboration in 2003, there are still problems with the quality and biases in patient decision aids.(3,5) Despite the recommended use of checklists, some patient decision aids are still biased towards low-value care and are not based on the best available evidence.(3) We followed the guidance from IPDAS and Elwyn et al. to produce a trustworthy tool that may improve customised care. We did this by following the three steps (evidence synthesis, patient experience, and patient-facing tool production).(37) Furthermore, we paid particular attention to including the most recent scientific evidence and presenting the pros and cons of different treatment strategies without recommending one treatment over another.

One of the aims of our tool was to decrease diagnostic uncertainty.(38) Adolescents may become confused and have difficulties understanding their condition if they sense diagnostic uncertainty among the clinician or do not understand the information provided.(39–42) From our experiences with the MAP-Knee Tool, we learned that developing targeted patient information is complex as explanations must be individualised and generalisable. To overcome this, we advise clinicians to use the tool as a guide and support tool but also include phrases and explanations tailored to each individual adolescent. However, to facilitate implementation in clinical practice where consultations need to be kept to a short time frame, the use of the MAP-Knee Tool must not extend the consultation time. Therefore, using standardised phrasings that the tool provides may ensure that the clinician covers important domains without using more time than they usually would during a consultation.

We were limited due to the lack of high-quality trials to support advice and the effectiveness of treatment strategies.(43) Despite our best efforts to include only high-quality evidence, we were limited by the number of randomised trials and systematic reviews conducted on non-traumatic adolescent knee pain. For example, we were only able to include a single narrative review concerning Sinding-Larsen-Johansson, whereas we included five systematic and narrative reviews concerning Osgood-Schlatter disease. Future improvement of the option grid of the MAP-Knee Tool would require a stronger research foundation. The MAP-Knee Tool should be a constantly evolving tool by making revisions when relevant research regarding treatment strategies has been conducted. Similar efforts have been made by performing living systematic reviews in Patellofemoral Pain and Achilles Tendinopathy that provide an up-to-date overview of evidence-based treatments.(44,45) For now, the MAP-Knee Tool is a physical tool, but to ease future revisions, it may be preferable to convert it into an online version. Still, it would be important also to use written materials such as a leaflet adolescents may bring home with them as such has been shown to assist patients with chronic pain in reconceptualising pain and reducing pain catastrophising.(46) Therefore, the MAP-Knee Tool should not be considered a tool that is only being used during the consultation but as a tool that extends beyond the clinical setting by using the leaflet. This can further substantiate the decrease in diagnostic uncertainty and the increased feeling of being validated as per the logic model.

The study has limitations. First, we only used the MAP-Knee Tool for Osgood-Schlatter disease during the end-user testing to limit time consumption. Despite the many commonalities between the tools developed for the different diagnoses, there may be some readability issues within the other tools that we did not capture. Second, the ability to present the pros and cons of the treatment strategies in the option grid relies on the scientific evidence available, which is generally inadequate for some of the diagnoses. Third, although the Argumentative Delphi process was conducted with international experts, we do not know how the tool will work in different social and cultural contexts as it was developed and user-tested in Denmark. Furthermore, we decided by consensus how to sub-group the treatment strategies, which may have inadvertently biased the presentation towards exercise-focused and not passive treatment strategies due to the research on exercise, which we have primarily in our group. Nevertheless, we were aware of this and did what we could to be as objective as possible.

One of the study’s main strengths is its systematic approach and use of end-user testing to ensure that the tool included relevant content and that the explanations and phrasings were understandable, readable, and matched the health literacy of the target group. Another strength is that we included clinicians and researchers with different backgrounds (e.g., medical doctors, psychologists, and physiotherapists) in the Argumentative Delphi process and medical doctors from various settings in the end-user testing. This may help the future implementation of the tool across different sectors as clinicians may find it valuable and meaningful regardless of which sector they are working in. This could ultimately lead to a higher degree of consistency within the clinical pathway for adolescents, which has recently been highlighted as being important by Ecclestone et al..(47)

### 4.2. Conclusion

Through a multi-step iterative process including systematic literature searches, an Argumentative Delphi process with international experts, and end-user testing, we developed a decision support tool for clinicians and adolescents with non-traumatic knee pain to support the entire consultation in clinical practice.

### 4.3. Practice Implications

The MAP-Knee Tool can potentially support clinicians and adolescents with non-traumatic knee pain in the clinical setting and ultimately lead to improved long-term outcomes for adolescents. The tool targets clinicians and adolescents with four components that may decrease diagnostic uncertainty and increase shared decision-making. Before future implementation of the tool in clinical practice, it is important first to investigate its feasibility in the clinical setting and then investigate if using the MAP-Knee Tool is more effective than usual practice in improving outcomes.

## Data Availability

All data produced in the present study are available upon reasonable request to the authors.

